# Uncovering the gastrointestinal passage, intestinal epithelial cellular uptake and AGO2 loading of milk miRNAs in neonates using xenomiRs as tracers

**DOI:** 10.1101/2021.08.24.21262525

**Authors:** Patrick Philipp Weil, Susanna Reincke, Christian Alexander Hirsch, Federica Giachero, Malik Aydin, Jonas Scholz, Franziska Jönsson, Claudia Hagedorn, Duc Ninh Nguyen, Thomas Thymann, Anton Pembaur, Valerie Orth, Victoria Wünsche, Ping-Ping Jiang, Stefan Wirth, Andreas C. W. Jenke, Per Torp Sangild, Florian Kreppel, Jan Postberg

## Abstract

**Background:** Human breast milk has a high microRNA (miRNA) content. It remains unknown whether and how milk miRNAs might affect intestinal gene regulation and homeostasis of the developing microbiome after initiation of enteral nutrition. However, this requires that relevant milk miRNA amounts survive gastrointestinal passage, are taken up by cells, and become available to the RNA interference (RNAi) machinery. It seems important to dissect the fate of these miRNAs after oral ingestion and gastrointestinal passage.

**Objective:** Our goal was to analyze the potential transmissibility of milk miRNAs via the gastrointestinal system in neonate humans and a porcine model *in vivo* to contribute to the discussion whether milk miRNAs could influence gene regulation in neonates and thus might vertically transmit developmental relevant signals.

**Design:** We performed cross-species profiling of miRNAs via deep-sequencing and utilized dietary xenobiotic taxon-specific milk miRNA (xenomiRs) as tracers in human and porcine neonates, followed by functional studies in primary human fetal intestinal epithelial cells (HIEC-6) using Ad5-mediated miRNA-gene transfer.

**Results:** Mammals share many milk miRNAs yet exhibit taxon-specific miRNA fingerprints. We traced bovine-specific miRNAs from formula-nutrition in human preterm stool and 9 days after onset of enteral feeding in intestinal cells of preterm piglets. Thereafter, several xenomiRs accumulated in the intestinal cells. Moreover, few hours after introducing enteral feeding in preterm piglets with supplemented reporter miRNAs (cel-miR-39-5p/-3p), we observed their enrichment in blood serum and in AGO2-immunocomplexes from intestinal biopsies.

**Conclusions:** Milk-derived miRNAs survived gastrointestinal passage in human and porcine neonates. Bovine-specific miRNAs accumulated in intestinal cells of preterm piglets after enteral feeding with bovine colostrum/formula. In piglets, colostrum supplementation with cel-miR-39-5p/-3p resulted in increased blood levels of cel-miR-39-3p and argonaute RISC catalytic component 2 (AGO2) loading in intestinal cells. This suggests the possibility of vertical transmission of miRNA signaling from milk through the neonatal digestive tract.

## Introduction

Milk from other species has been a part of human nutrition since the Neolithic period, and has left traces in the genomes of many modern humans. European hunter-gatherers were unable to digest lactose after weaning, but this changed with the migration of Neolithic populations who practiced dairy farming and exhibited genetic adaptations associated with lactase (LCT) persistence after infancy. This led to the dominance of lactase persistence markers in people of Caucasian descent today. (1, 2). Similar genetic adaptations have been observed in African and Middle Eastern populations. (3–5). Apparently, domestication of mammals was associated with the positive-selection of genetic traits allowing a beneficial niche construction, whereby adult carriers could exploit xenobiotic milk as additional food source (6–10). Notably, nowadays bovine milk in particular plays a global role on the world market as fresh milk or in processed products far beyond industrial baby milk products (11).

Genetic markers aside, LCT persistence and non-persistence are phenomena of gene regulation (2). In preterm neonates, LCT activity increases after beginning enteral feeding, with formula-fed infants exhibiting lower activity levels than milk-fed infants (12). Beyond LCT, a neonate piglet model study found that formula feeding resulted in different intestinal gene regulation compared to colostrum feeding or total parental nutrition. This is significant because the type of feeding can impact the risk of necrotizing enterocolitis (NEC) in premature infants. Infants who receive bovine milk-based formula have a higher risk of NEC compared to those who are fed breast milk or colostrum (13–16). However, the underlying molecular mechanisms responsible for nutrition-dependent differences in gene regulation remain unclear. Differences in post-transcriptional gene regulation through RNA interference (RNAi) could hypothetically be due to differences in the miRNA profiles of human milk and formula, as many formulas contain bovine milk. This post-transcriptional gene silencing (PTGS) can influence gene regulatory networks (Supplemental Figure 1). In PTGS, microRNAs (miRNAs) interacting with Argonaute (Ago) protein family members target specific mRNAs via base-pairing of their 5’-seed (17–19). Perfect or near-perfect base-pairing promotes the decay or translational repression of targeted mRNAs (20). MiRNAs occur in cells and ubiquitously in body fluids, among which human breast milk seems to exhibit the highest total amount (21). Many processed milk-products and formulas contain miRNAs (22). MiRNAs are known to facilitate communication between cells. In this regard, it is widely accepted that these miRNAs may be packaged into extracellular vesicles (EVs), such as exosomes. However, miRNAs can also occur in vesicle-free forms (23).

There is an ongoing debate in science and society about possible adverse effects of milk consumption with particular emphasis on bovine milk and in some instances focusing the controversy on its miRNAs content (24–26). This discussion lacks comparative and experimental data on the biological significance of milk miRNAs. Contradictory results concerning the transmissibility of milk miRNAs have been published by different groups recently, who did not see evidence for milk miRNA uptake in newborn mice (27) or, respectively, demonstrated miRNA internalization in human intestinal cells *in vitro* after simulating infant gut digestion of milk (28). Interestingly, oral administration of cow milk exosomes in mice resulted in their apparent bioavailability, albeit low compared to intravenous administration (29). There remains thus an urgent need for further rigorous experimental data to inspire the debate and address the open problems: Are milk miRNAs functional, or are they a nucleo(t/s)ide source, only? Although several studies addressed this problem recently (30–32), we currently neither know well whether milk miRNAs build a path of vertical signal transmission from mother to the new-born nor whether interferences of xenomiRs with human target mRNAs can influence developmental pathways (33).

In this study, we aim to examine the transmissibility of milk miRNAs through the digestive system in neonates and a porcine model, to shed light on their potential impact on gene regulation and vertical transmission of developmental signals.

## Methods

### Clinical trial registration

All analyses including those using human fecal specimens were considered exploratory. Clinical trial registration was not required since participants were not prospectively assigned to an intervention, the research was not designed to evaluate the effect of an intervention on the participants, and no effect was being evaluated in an health-related biomedical or behavioral outcome.

### Samples and animal procedures

Human milk (<6-72 hrs post partum) and stool specimens were collected with approval of the Witten/Herdecke University Ethics board (No. 41/2018) after informed written consent was obtained by all involved donors. Animal milk was purchased or collected with the help of Wuppertal Zoo, local farmers, or veterinary medics. A participant flowchart for human and animal milk specimen collection is provided (Supplemental Figure 2). All animal procedures were approved by the National Council on Animal Experimentation in Denmark (protocol number 2012-15-293400193). Piglet (landrace × large white × duroc; delivered by caesarean section on day 105 [90%] of gestation) blood and gut biopsies derived from an ongoing experimental series described previously (34). A participant flowchart for piglet intestinal specimen collection is provided (Supplemental Figure 3). Reported milk nucleic acids concentration were used as guideline for reporter miRNA supplementation of bovine colostrum bolus, which was intended for piglet feeding (23 mg/L ± 19 mg/L [8.6-71 mg/L]) (35). We supplemented bovine colostrum bolus with RNA oligonucleotides mimicking both putatively processed strands of *Caenorhabditis elegans* (cel), cel-miR-39-5p and cel-miR-39-3p, and adjusted their amount to achieve approx. 5-10% of human milk’s total nucleic acids content. Before the feeding experiments, we checked whether the spiked cel-miRNAs became enriched within the EV fraction of human, bovine and porcine colostrum as described below. Three neonatal low-birth-weight piglets from the same litter (mean birth weight 1,119 g [1,071-1,167 g]; normal birth weight approx. 1,400-1,500 g) received initial feeding (t_0_) of bovine colostrum bolus (15 mL/kg) enriched with 39 g/L casein glycomacropeptide (CGMP), 2.8 g/L osteopontin (OPN) and a mix of cel-miR-39-5p/-3p reporter miRNAs. As control, three other piglets from the same litter received colostrum bolus without reporter miRNA supplementation (mean birth weight 1,047 g [862-1,240 g]. After 2.5 h, the piglets received another bolus to facilitate gut motility. At different time points post-feeding (t_1_=30 min, t_2_=1 h, t_3_=2 h, t_4_=3 h, t_5_=5 h and t_6_=7 h) 1 mL peripheral blood was sampled. Total RNA was used for miRNA library preparation and successive analyses of enrichment of both reporter cel-miRNAs by qPCR, whereas confirmatory miRNA-seq was conducted 7 h post-feeding only.

Enrichment of piglet intestinal epithelial cells from intestinal mucosa biopsies was done as follows. Pigs were euthanized using 200 mg/kg intraarterial sodium pentobarbital after anesthesia administration. The GI tract was immediately removed, and the small intestine was carefully emptied of its contents. 30 cm of the distal small intestine were used for cell collection. The segment was flushed with 10 mL of pre-chilled NaCl 0.9% (Fresenius Kabi, Uppsala, Sweden) and then cut open along the length. The upper layer of the intestinal mucosa was gently scraped off with a microscopic slide and transferred in a 15 mL tube containing 12.5 mL PBS containing FBS (final concentration 1.12 µL/mL). Homogenization was achieved by pipetting with a plastic Pasteur pipette for about 1 min and then stirring the solution at 500 rpm for 20 min at room temperature. The homogenized suspension was filtered through a 70 µm cell sieve and spun down at 500x g at 4°C for 3 min. The cell pellet was resuspended in 5 mL modified PBS (see above), filtered and centrifuged again. The pellet was then resuspended in 1 mL red blood cell lysis buffer (1x, InvitrogenTM, ThermoFisher Scientific), incubated for 5 min at room temperature and then spun down at 500x g at 4°C for 3 min. The cell pellet was washed again and finally resuspended in 1 mL modified PBS (see above). From the resulting single cell suspension 250 µL were used for RNA purification. The authors state that they have obtained appropriate institutional review board approval or have followed the principles outlined in the Declaration of Helsinki for all human or animal experimental investigations.

### Milk extracellular vesicle (EV) fractionation and detection of supplemented cel-miR

Human, bovine and porcine sera obtained by 2x 10 min centrifugation at 3,000x g were spiked with various concentrations of cel-mir-39-5p/3p (final concentrations: 5 µM, 1 µM, 0.2 µM or 40 nM) followed by overnight incubation at 4°C prior to EV enrichment as described elsewhere (36). Briefly, ExoQuick exosome precipitation reagent (SBI System Biosciences) was added upon manufacturer’s recommendation followed by another overnight incubation at 4°C. EVs and supernatant were separated at 13,000x g for 30 minutes at 4°C. Then RNA was purified as described below. RNA was analyzed on an Agilent Bioanalyzer 2100 and by polyacrylamide gel electrophoresis (PAGE). Endogenous controls (miR-148a-3p, miR-143-3p, miR-99b-5p, and U6 snRNA) and spiked cel-miR-39-5p/3p were assessed using the Mir-X™ miRNA First Strand Synthesis Kit (Takara) in combination with qPCR and primers as listed (Supplemental Table 1A).

### Nucleic acids

Total RNA was purified from milk serum, blood serum, radically grinded tissue, cells and stool samples using Trizol reagent (Sigma, MO, USA) upon manufacturer’s recommendations followed by isopropanol precipitation and dilution in water. RNA quality and quantity were assessed by microcapillary electrophoresis using an Agilent Bioanalyzer 2100 similarly as described (37). A list of oligonucleotides used is provided as appendix (Supplemental Table 1A).

### Selection of cattle-specific reporter miRNAs

For deeper investigation of their putative influence in a human host we selected several bta-miRs using relative quantities and the validated the absence of similar miRNAs in the human miRNome as the main selection criteria. Selected bta-miRNA quantities were estimated via their ranks (#n) within *Bos taurus* total miRNAs: bta-mir-2904 (#19); bta-mir-2887 (#67); bta-mir-2478 (#93); bta-mir-3533 (#141); bta-mir-2428 (#169); bta-mir-2440 (#172); bta-mir-677 (#191); bta-mir-2892 (#229); bta-mir-2404 (#232); bta-mir-2361 (#237); bta-mir-2372 (#278); bta-mir-2304 (#284). Typically, human mRNA target prediction for those bta-miRNAs via miRDB (38) revealed dozens of potential high scoring 3’-UTR targets, with transcripts of several master regulatory genes among them (Supplemental Table 2). For example, for bta-miR-677-5p (5’-C**<u>UCACUGA</u>**UGAGCAGCUUCUGAC-3’, probable seed-sequence underlined), transcription factor SP4, *zink finger and homeobox family member 1* (ZHX1) and the chromatin remodeler ATRX were among the predicted human mRNA targets (n=655). Importantly, such algorithms usually deliver large numbers of false-positive results, which might be invalid for specific cell types-of-interest. But before interrogating this problem, it is crucial to address whether (xenobiotic) miRNAs stay intact during the gastric and intestinal transit and eventually do reach potential mRNA targets within cells.

### Real time PCR

Quantitative measurement of miRNAs was analyzed from cDNA libraries (as described in (37)) using primer pairs as listed (Supplemental Table 1A) and the QuantiTect SYBR Green PCR Kit (Qiagen) using a Corbett Rotor-Gene 6000 qPCR machine. Specifically, a combination of miR-specific forward primers [bta/pan-miR-*name*-5p/-3p-fw] and library-specific reverse primers [library_P*n*_miR_rv]) was used (Supplemental Table 1B). PCR conditions were as follows: 95°C for 15 min, 40 cycles of (95°C for 15 s, 60°C for 30 s). Amplicon melting was performed using a temperature gradient from 55–95°C, rising in 0.5°C increments. For relative comparative quantification of quantitative expression fold changes we utilized the ΔΔCt method (39) using stable miR-143-3p, miR-99b-5p and let7b-5p for normalization.

### Deep sequencing of miRNAs (miRNA-seq) and bioinformatics pipeline

Small RNAs (18-36 nt) were purified from total RNA using polyacrylamide gel electrophoresis (PAGE). The subsequent cDNA library preparation utilizes a four-step process starting with the ligation of a DNA oligonucleotide to the 3’-end of the selected small RNAs, followed by ligation of a RNA oligonucleotide to the 5’-end of the selected small RNAs. The resulting molecules were transcribed via reverse transcription (RT) and amplified via PCR (37). Subsequently, after final quality checks by microcapillary electrophoresis and qPCR, the libraries were sequenced on an Illumina HiSeq 2000 platform (single end, 50 bp). This work has benefited from the facilities and expertise of the high throughput sequencing core facility of I2BC (Research Center of GIF – http://www.i2bc.paris-saclay.fr/). The initial data analysis pipeline was as follows: CASAVA-1.8.2 was used for demultiplexing, Fastqc 0.10.1 for read quality assessment and Cutadapt-1.3 for adaptor trimming, resulting in sequencing reads and the corresponding base-call qualities as FASTQ files. File conversions, filtering and sorting as well as mapping, were done using ‘Galaxy’ (40–42), a platform for data intensive biomedical research (https ://usegalaxy.org/), or ‘Chimira‘ (43), sRNAtoolbox (44) or ‘miRBase‘ (45), respectively. For selected bta-mir we used the miRDB custom option for sequence-based prediction of potential human mRNA targets (38). miRDB’s main criterion to report a potential miRNA:mRNA interaction is based on perfect or near-perfect base pairing of the 5’-seed region of the miRNA with the 3’-UTR of an mRNA. The presence of multiple potential targets in a 3’-UTR is incorporated into the ranking of potential targets.

### Argonaute co-immunoprecipitation (Co-IP)

Association of reporter cel-miR-39 co-fed with milk with Argonaute (AGO2) in piglet intestinal biopsies was analyzed using co-immunoprecipitation and qPCR. For native pan-AGO2 precipitation from intestinal tissue we used monoclonal mouse anti-pan-AGO2 antibodies (Sigma-Aldrich, MABE56, clone 2A8) similarly as described previously (46). This antibody exhibits cross-species reactivity with AGO2 from mouse, humans, and most probably from other mammals as well. After final blood sampling 7-7.5 h post-feeding proximal and distal tissues were taken at euthanasia, deep frozen at -20°C and shipped on dry ice. Prior to immunoprecipitation piglet intestinal tissue from a liquid nitrogen storage tank was powdered using a Cellcrusher™ (Cellcrusher, Co. Cork, Ireland) pre-chilled in liquid nitrogen. Subsequently, intestinal tissue powder was suspended in lysis buffer (20 mM Tris-HCl, pH7.5, 200 mM NaCl, 2.5 mM MgCl_2_, 0.5% Triton X-100). Samples were incubated overnight with 3 μg anti-pan-Ago (2A8) and 25 μL magnetic protein A beads (Diagenode). Following the enrichment of immunocomplexes on a magnetic rack and several washes with PBS, the samples were heated to 95°C for 15 min, and then RNA was purified from the immunocomplexes using Trizol and isopropanol precipitation. Then RNA converted into cDNA libraries and cel-miR-39-5p/-3p were analyzed from cDNA libraries by qPCR.

### Luciferase assay

We performed this experiment to rule out exemplarily that the observed bta-miR-candidates could be simply miRNA-like sequence artefacts without any function, we interrogated whether bta-miR-677, a candidate miRNA with intermediate abundance in milk can silence luciferase expression *in vitro* via a predicted 3’-UTR target sequence in a human HeLa cell-based assay. Therefore, a Sac1/Nhe1 fragment with sticky ends was hybridized from oligonucleotides through incubation in a boiling water bath and successive chilling to room temperature (Supplemental Table 1B). Then we cloned the fragment containing a 7 bp 3’–UTR target motif reverse complementary to the 5’-seed (nucleotides 2-8) of bta-miR-677-5p into the 3’-UTR region of the firefly luciferase gene (luc2) encoded in the pmirGLO dual luciferase expression vector (Promega) (Supplemental Figure 4A). For bioluminescence calibration this vector contained a 2^nd^ luciferase cassette (*Renilla*; hRluc-neo fusion). Subsequently, pmirGLO was co-transfected into HeLa cells using Lipofectamin 2000 (ThermoFisher) with bta-mir-677-like hairpin RNAs or non-luciferase directed controls, followed by bioluminescence quantification using a GloMax® Multi Detection System (Promega). We observed very robust silencing of firefly luciferase upon bta-mir-677-like co-transfection, suggesting that at least bta-miR-677-5p can function as valid miRNA in human cells (Supplemental Figure 4B).

### Insertion of microRNA-hairpins into Ad5 vectors

To assess the effects of miRNA overexpression in cell culture Ad5-based replication deficient ΔE1 vectors harboring an hCMV promoter-driven expression cassette for the concomitant expression of EGFP (as transduction marker) and miRNAs were generated based on the ‘BLOCK-iT Pol II miR RNAi Expression Vector Kits’ protocol (Invitrogen). Vectors were serially amplified on 293 cells and purified by CsCl density centrifugation. The vector titers were determined by OD260 and vector genome integrity was confirmed by sequencing and restriction digest analysis (47). Hairpins mimicking bovine pre-mirs were designed to express miRNAs, which corresponded to bta-miR-677-5p, bta-miR-2440-3p, bta-miR-2361-5p. Details of vector design can be obtained upon request. The expression of bta-mir-677-like hairpins, bta-miR-677-5p and bta miR-677-3p, respectively, was evaluated by qPCR using the Mir-X™ miRNA First Strand Synthesis Kit (Takara).

### Cell culture and Ad5 transduction

Human intestinal epithelial cells (HIEC-6 [ATCC CRL-3266]) cells were cultured upon manufacturer’s recommendations. Flow cytometric analyses of HIEC-6 cells transduced with different ranges of multiplicity of infection (MOI) were used to define a transduction efficiency and to determine the optimal MOI (Supplemental Figure 5). Considering transduction efficiency and compatibility, we subsequently used MOI 10,000 in experimental routine. One day pre-transduction, 10^6^ HIEC-6 cells were seeded in 6-well cell culture dishes. The following day, cells were washed with PBS, provided with fresh medium and subsequently transduced with respective Ad5 vector particles. Cells were harvested 96 h post-transduction. The expression of bta-mir-677-like hairpins, bta-miR-677-5p and bta-miR-677-3p, respectively, was validated using the Mir-X™ miRNA First Strand Synthesis Kit (Takara) in combination with qPCR. The results confirmed the expression of bta-mir-677-like hairpins, bta-miR-677-5p, but a bias for bta-miR-677-3p suggesting preferential processing of the 5-p guide strand (Supplemental Figure 6). For Western blot analyses cell pellets were homogenized in RIPA buffer (50 mM Tris HCl pH 8.0, 150 mM NaCl, 1% NP-40, 0,5% sodium deoxycholate, 0,1% SDS plus cOmplete^™^ Mini Protease Inhibitor Cocktail [Sigma-Aldrich]). Alternatively, cells were grown on coverslips for immunofluorescence microscopy. Here, after Ad5-mediated miRNA transfer successful expression of the miRNA transgene could be indirectly confirmed using immunofluorescence microscopy and flow cytometry for detection of green fluorescent protein (GFP) that was co-expressed with bta-mir-677-like hairpins from the same transcript.

### Western blot analyses

Homogenized pellets were mixed with SDS. Proteins were separated by SDS-PAGE using 5% stocking and 8% running gels and subsequently transferred to a nitrocellulose blotting membrane (Amersham Protran Premium 0.45 μm NC). Membranes were blocked 1% BSA in TBST (20 mM Tris;150 mM NaCl; 0.1% Tween 20; pH 7.4). Polyclonal rabbit anti-ATRX (Abcam; ab97508), polyclonal rabbit anti-DDX3 (Abcam; ab235940), polyclonal rabbit anti-ZHX1 (Abcam; ab19356), polyclonal rabbit anti-PLCXD3 (Sigma-Aldrich; HPA046849), polyclonal rabbit anti-GAPDH (Sigma-Aldrich; G8795) were used as primary antibodies at 1:500 dilution in blocking buffer, except anti-GAPDH (dilution 1:20,000 in blocking buffer). For internal normalization anti-GAPDH was used in parallel and simultaneously with each other primary antibody. Secondary antibodies were goat anti-rabbit IRDye 800CW (LI-COR; 926-32211) and goat anti-mouse IRDye 680RD (LI-COR; 926-68070), each being diluted 1:10,000 as a mix in blocking buffer. Fluorescence was monitored using the Odyssey CLx Imaging system (LI-COR) with the 2-color detection option and analyzed using Image Studio software (LI-COR).

### Immunofluorescence microscopy

5x 10^5 HIEC-6 cells were grown on coverslips in 6-well plates and transduced as described above. 96 h post-transduction cells were fixed for 10 min in 2% PFA/DPBS, washed twice with DPBS and then permeabilized using 0.5% Triton X-100/DPBS, followed by 0.1N HCl for exactly 5 min for antigen retrieval and successive washes with DPBS. Blocking was done in 3% BSA/0.1% Triton X100/DPBS. Subsequently, the primary antibodies were incubated in blocking buffer for 1 h/37°C: 1. polyclonal rabbit anti-ATRX (Abcam; ab97508) at 1:100, polyclonal rabbit anti-DDX3 (Abcam; ab235940) at 1:500, polyclonal mouse anti-ZHX1 (Abcam; ab168522) at 1:100. Then polyclonal goat anti-rabbit Cy3 (Jackson ImmunoResearch) or polyclonal goat anti-mouse Cy3 (Jackson ImmunoResearch) were used as secondary antibodies, each at 1:100. Eventually, 4’,6-diamidino-2-phenylindole (DAPI) was used for DNA counterstaining at 0.1 μg/mL followed by washes and mounting with Prolong Gold (Invitrogen). Acquisition of images was done with a Nikon Eclipse Ti Series microscope. Fluorochrome image series were acquired sequentially generating 8-bit grayscale images. The 8-bit grayscale single channel images were overlaid to an RGB image assigning a false color to each channel using open source software ImageJ (NIH Image, U.S. National Institutes of Health, Bethesda, Maryland, USA, https://imagej.nih.gov/ij/).

### Deep sequencing of messenger RNAs (mRNA-seq) and bioinformatics pipeline

Total RNA was purified from Ad5-transduced HIEC-6 cells expressing a nonsense-miR or bta-miRs using Trizol reagent (Sigma, MO, USA) upon manufacturer’s recommendations. RNA quality and quantity were assessed by microcapillary electrophoresis using an Agilent Bioanalyzer 2100. All specimens had a RIN between 8.9 and 9.2 and were subsequently used for sequencing library preparation. Enrichment of mRNAs was done using the NEBNext Poly(A) mRNA Magnetic Isolation Module (New England BioLabs, #E7490S) upon manufacturers’ recommendations. Subsequently cDNA libraries were prepared using the NEBNext Ultra II DNA Library Prep Kit for Illumina (New England BioLabs, #E7645S) upon manufacturers’ recommendations in combination with NEBNext Multiplex Oligos for Illumina (Index Primer Set 1) (New England BioLabs, #E7335G). Before multiplexing, library quality and quantity were assessed by microcapillary electrophoresis using an Agilent Bioanalyzer 2100. Deep sequencing was done using an Illumina HiSeq2500. The initial data analysis pipeline was as follows: CASAVA-1.8.2 was used for demultiplexing, Fastqc 0.10.1 for read quality assessment and Cutadapt-1.3 for adaptor trimming, resulting in sequencing reads and the corresponding base-call qualities as FASTQ files. File conversions, filtering and sorting as well as mapping, were done using ‘Galaxy’ (40–42) (https ://usegalaxy.org/), serially shepherding the FASTQ files through the following pipeline of tools: 1. FASTQ groomer (Input: Sanger & Illumina 1.8+ FASTQ quality score type) (48); 2. TopHat for gapped-read mapping of RNA-seq data using the short read aligner Bowtie2 (Options: single end, *Homo sapiens*: hg38) resulting in a BAM formatted accepted hits file (49). 3. featureCounts to create tabular text file (Output format: Gene-ID “\t” read-count [MultiQC/DESeq2/edgeR/limma-voom compatible]). 4. Prior to DESeq2 normalization, a tabular file containing catalogued columns of read counts for all experiments was arranged in Microsoft Excel for Mac 16.32. and then uploaded as a tabular text file to Galaxy. 5. annotadeMyIDs was used to assign gene symbols and gene names to Entrez IDs.

### Biostatistical tests

Biostatistical tests were carried out using GraphPad Prism 8.4.3 software. By multiple comparisons we studied the differential occurrence of detected milk miRNAs between hominidae vs. bovidae, hominidae vs. suinae, bovidae vs. suinae as well as omnivores vs. herbivores, omnivores vs. carnivores, or herbivores vs. carnivores using DESeq2-normalized miRNA data. Normal distribution tests were conducted for each miRNA assuming alpha=0.05 (accepted significance level) and using Shapiro-Wilk test or, alternatively, Kolmogorov-Smirnov test with the result that the majority of all miRNAs were normally distributed across all groups. For group-wise comparisons and to identify differentially distributed miRNAs, we conducted 2-way ANOVA test. Significance level accepted was alpha=0.05. Tukey’s multiple comparisons test was selected as a method to correct for multiple comparisons and to report multiplicity adjusted P value for each comparison.

Differential effects of selected Ad5-mediated bta-miR expression were studied in HIEC-6 in comparison to HIEC-6 expressing a Ad5-mediated nonsense-miR (control group). Relative fold-changes were compared between the whole HIEC-6 transcriptome (mRNA) and the lists of expressed predicted targets. Statistical power calculations were done using the nonparametric Wilcoxon–Mann–Whitney method.

### Statement on methodological limitations

We identified some methodological limitations that should be considered when evaluating our results: An important limitation concerns the interpretation of the spike experiments, especially the results of fractionation EVs. We used the ExoQuick precipitation method to gain initial insight into whether a miRNA added to colostrum is associated with the EV fraction in a mechanistically unknown manner. This method is classified as a ’high recovery, low specificity’ method according to the ’Minimal information for studies of extracellular vesicles 2018’ (50), which does not allow high purity separation of the different subtypes of EVs. Importantly, future functional studies on the role of milk miRNAs in vertical transmission of maternal miRNA signals should therefore have higher quality requirements regarding EV purification and controls, following MISEV2018 guidelines.

The study used a small sample (only three piglets per group), which may not be sufficient to represent the entire population. The use of animal studies may also limit the generalizability of the results to humans. The amount of RNA oligonucleotides used for supplementation was based on a single study and may not reflect the typical content of nucleic acids in human or animal milk. The sequencing method used, and the bioinformatics pipeline may also introduce bias or error in data analysis. Confirmatory miRNA sequencing was performed only 7 hours after feeding, which limits the ability to assess the overall time course of reporter miRNAs in piglets. The process of enrichment of piglet intestinal epithelial cells may lead to bias or loss of important information during the cell collection and purification steps. These limitations highlight the need for caution in interpreting the results and suggest that further studies with larger sample sizes and more comprehensive sampling and sequencing methods are needed to confirm the results.

## Results

### Mammalian milks shared many miRNAs yet exhibited species-specific miRNA fingerprints

We used miRNA deep sequencing (miRNA-seq) to determine miRNA profiles of human (hsa), cattle (bta), goat (cae), and sheep (ogm) milks. In addition, we speculated that an evolutionary context of milk miRNA profile differences might exist, for example between omnivores, herbivores, and carnivores. Therefore, our study also included milk from pig (ssc), okapi (ojo), horse (eca), donkey (eas), cat (fca), and dog (clu) (Figure 1A).

**Figure 1.**
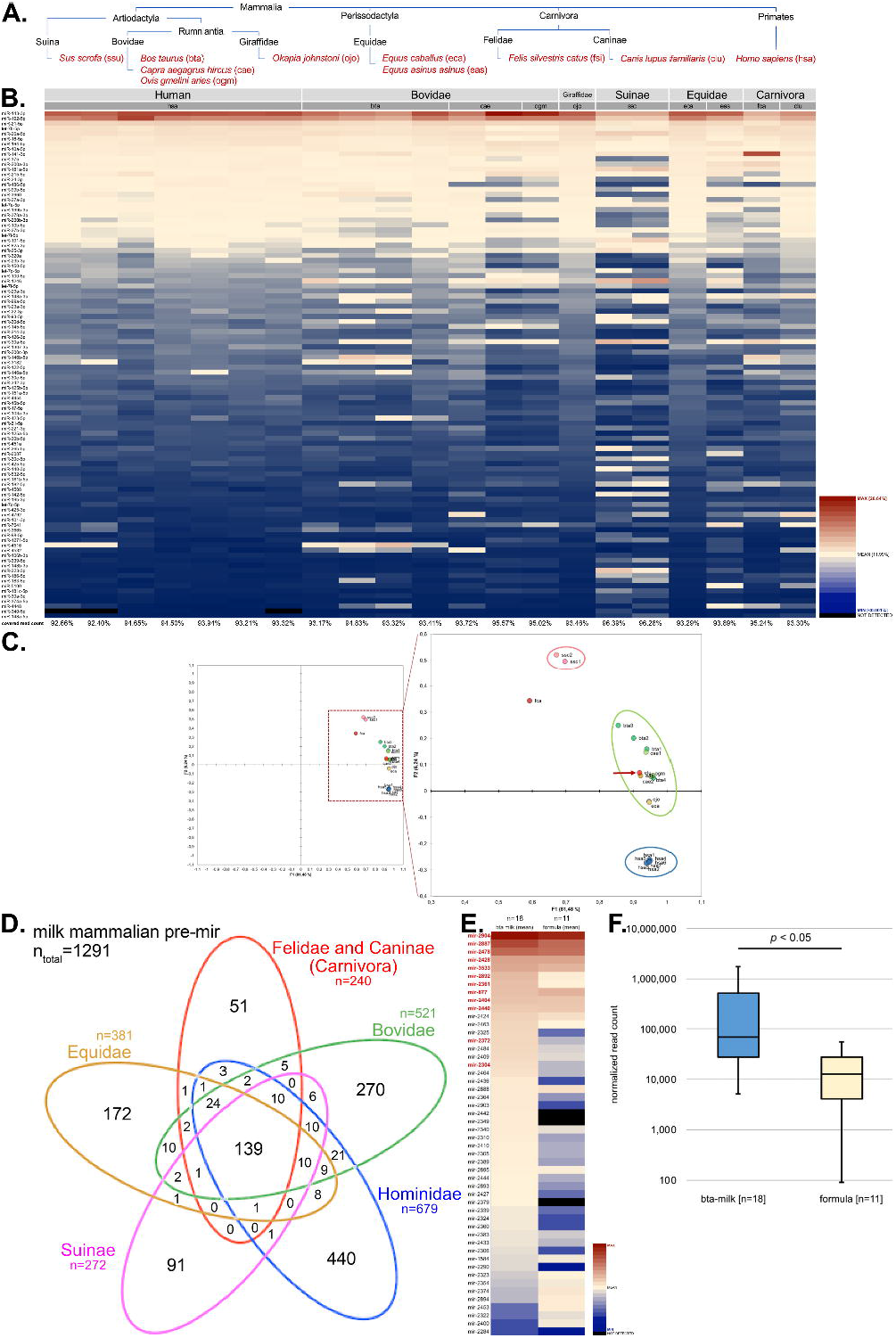
Comparative profiling of mammalian milk miRNAs. **A.** Phylogenetic tree of specimens under analysis (red font), which included mostly domesticated species with the exception of okapi (ojo) and human (hsa). Additionally, we determined miRNA profiles from yak cheese and water buffalo cheese (light red font), which are part of the available dataset, but are not part of the below analyses due to insufficient total read counts and quality. **B.** Heat map of miRNAs conserved in all taxa analyzed. Only the top 100 miRNAs found in all species are shown robustly representing over 90% of the total read count in all specimens (see values below each column). A color gradient was used for the visualization of each miRNA’s abundance (read counts per million; CPM) in each specimen. MicroRNAs were sorted in descending order with reference to the median CPM of human specimens. **C.** Results of principal component analyses of microRNAs shared by all taxa under investigation. Quadrants I and IV (section magnified on the right) harbor all specimens whereby all human specimens (blue: hsa1-7) are clustering. Slightly separated is a separated scattered cluster harboring all herbivorous specimens (yellow to green: bta1-4, cae1-2, eas, eca, ogm, ojo) and one dog specimen (red arrow: clu). Above is a mini-cluster comprising both porcine specimens (pink: ssc1-2) and separated the cat specimen (red: fca). **D.** Venn diagram showing that the numbers of pre-miRNAs (pre-mir) shared by all or some taxa or, respectively, which were identified being taxon-specific. For this analysis pre-mir counts were used rather than miR-counts to counteract a potential differential taxon-specific 5p-/3p-dominant processing. **E.** Heat map of the top 50 *Bos taurus* (bta)-specific pre-mir in raw and industrially processed milk specimens (n=18) in comparison with powdered milk/formula product specimens (n=11). The median CPM (mapped reads count per million) was used as sorting criterion in descending order. Red font: miRNA was used for human mRNA target prediction via the sequence-based custom prediction option of miRDB (38). Cave! Bovidae in heat map include goat and sheep, whereas ranks were determined from cattle only. **F.** Quantitative analysis of total miRNA read counts in raw and industrially processed milk specimens (n=18) in comparison with powdered milk/formula product specimens (n=11).

We detected 1291 hairpin-like pre-miRNAs (pre-mir) across all taxa. 139 of these pre-mirs occurred in all taxa. Notably, during the biogenesis of mature single-stranded miRNAs (miR), each single pre-mir can theoretically give rise to two mature miRs through processing of its guide (5p) and/or passenger (3p) strand. We visualized the top 100 milk miRNAs from 21 specimens as a heat map (Figure 1B) and as a dot plot (Supplemental Figure 7). In all specimens these top 100 miRNAs represented more than 90% of the total mapped miR read counts (range 92.40-96.39%).

The heat map and dot plot suggested a stable quantitative distribution of many miRNAs in the milk of most species, although principal component analysis revealed somewhat separate clusters in detail (Figure 1C): Cluster ‘blue’ contained all human specimens, cluster ‘green’ contained all herbivores as well as the dog, while the pig and cat appeared somewhat separated. For deeper investigation of the putative pairwise differences between individual hsa-miRs and bovid miRs, we used ANOVA tests (Supplemental Table 3). However, we only identified a few differences, with only miR-192-5p falling below p≤0.0001 and 3 miRs falling between 0.001≤p≤0.05 (miR-1246, miR-146b-5p, miR-4516). We also observed few differences, when comparing hsa-miRs vs. ssc-miRs, bovid miRs vs. ssc-miRs, omnivores miRs vs. herbivores miRs, omnivores miRs vs. carnivores miRs and herbivores miRs vs. carnivores miRs (Supplemental Table 1).

Apart from the many miRNAs shared between all species, we observed numerous miRNAs being apparently shared by some taxa only or being taxon-specific. We undertook a systematic search to identify taxon-specific pre-mir and to analyze their non-overlapping or partial overlapping in different taxa using mapped miRNAs for Blast searches and the curatated database miRbase as a source (45) (Supplemental Table 4). For each taxon, we identified several hundreds of milk pre-mirs (carnivores: n=240; bovids: n=521; human: n=679; pigs: n=272; equids: n=381), but only a fraction was shared between all or some taxa (Figure 1D). For example, 225 pre-mir were shared between humans and cattle. Importantly, 440 pre-mir were human-specific and 270 were bovids-specific. We found many more pre-mir being specific for other taxa (pigs [n=91], equids [n=172] and carnivores [n=240]). Next, we exploited taxon-specific dietary xenomiRs from foreign species’ milks as molecular tracers to study their gastrointestinal (GI) passage as well as systemic and cellular uptake in human or animal models.

### Dietary-derived bta-miRs survived the gastrointestinal transit in human newborns

The heat map in Figure 1E illustrates the 50 top-ranked bovid-specific pre-mirs (specimens: 8 cattle, 1 yak, 1 water buffalo) (Figure 1E; left column). Further, 11 formula/powdered milk products are shown (Figure 1E; right column) (compare Supplemental Table 1A). Most bovid-specific pre-mir from liquid milk were present in formula products with similar relative quantities, but mostly with lower total miRNA concentrations (Figure 1F). To study the GI passage of milk miRNAs taken in through the diet, we selected *Bos taurus* (bta)-specific (pre-) miRNAs as tracers. We defined their relative quantities and their validated absence in the human miRNome as main selection criteria (Figure 1E, red font). Subsequently, we examined a randomly selected preterm neonate stool sample collection from a neonatology intensive care unit. The donors were given routinely formula supplementation (mean gestational age: 29.2 weeks [24.4-35.7]) (51). Quantitative real-time PCR (qPCR) analyses were performed using cDNA libraries prepared from purified small RNAs and specific sets of bta-miR or pan-miR primer pairs (Supplemental Table 1B). As a matter of fact, we detected all interrogated bta-miRs intact in all pre-term stool samples (Figure 2A).

**Figure 2.**
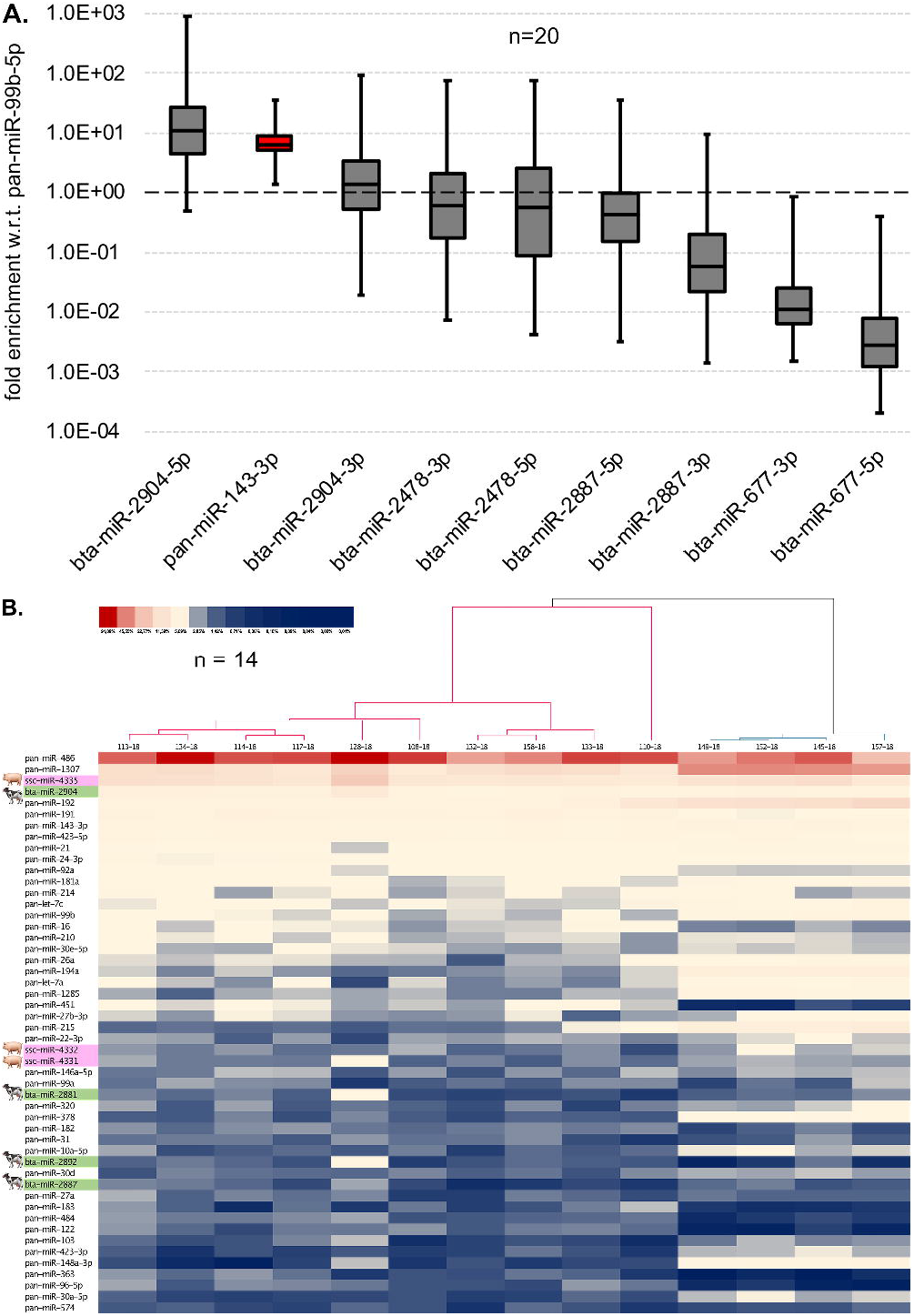
Xenobiotic bta-miRs survive the gastrointestinal passage in human preterm neonates, and in preterm piglets they become enriched in intestinal epithelial cells. **A.** This chart shows the enrichment of bovine (bta) miRNAs in infant stool when fed with formula. Results were obtained via qPCR for selected bta-miRs. For comparison, pan-miR-143-3p that is shared by humans and cattle is shown (red box, ‘pan’ prefix means that a miRNA is indistinguishable in humans and cattle). The relative enrichment was normalized with respect to pan-miR-99b-5p using the ΔΔCt method. For each PCR, we analyzed n=20 specimens with at least 3 replicated for each set of primers. **B.** Heat map of miRNAs detected in enriched porcine preterm neonatal intestinal cells (n=14 specimens) following 9 days of enteral bovine colostrum/formula supplementation. Only the top 50 miRNAs found in all species are shown. A color gradient was used for the visualization of each miRNA’s abundance (read counts per million; CPM) in each specimen. MicroRNAs were sorted in descending order with reference to the median CPM of all specimens. Prefix ‘pan’ for miRNA denotes that it is present in multiple species (here: pig and cattle)

### In preterm piglets cattle-specific miRNAs became enriched in intestinal cells after 9-days of enteral nutrition with bovine colostrum or formula supplementation

For deeper insights we used a preterm neonate piglet model to study the intestinal cellular uptake of milk miRNAs after prolonged feeding in vivo. This preterm neonate piglet model is being comprehensively used to study the effects of enteral feeding introduction post-delivery (13–16, 34). Therefore, we used cattle-specific colostrum- or formula-derived bta-miRs as tracers. For this experiment we prepared miRNA libraries for deep-sequencing from enriched intestinal epithelial cells 9 days after introducing enteral feeding and subsequent continuous supplementation with bovine colostrum or formula. Strikingly, we found that 4 bta-miRs were enriched within the 50 top-ranked intestinal cellular miRNAs (Figure 2B). Thereby, bta-miR-2904 occupied rank #4 suggesting that a dietary miRNA could possibly reach biologically significant concentration levels if being taken up by cells. Apparently, the amount of cellular uptake was influenced by the bta-miR concentration in bovine milk (bta-miR-2904: #4 in intestinal cells (IC) vs. #1 (bta-specific miRs) in bovine milk (BM); bta-miR-2881: #31/not ranked in IC/BM; bta-miR-2892: #37/6 in IC/BM; bta-miR-2887: #39/2 in IC/BM; compare Figure 1E).

Notably, we also tested the presence of bta-miRs in cerebrospinal fluid samples from piglets receiving enteral feeding of bovine colostrum (52), but we did not detect any.

### Feeding with cel-miR-39-5p/-3p-supplemented colostrum bolus led to rapid bloodstream enrichment of this reporter miRNA

Following the observation of xenomiRs enrichment in intestinal cells of neonate piglets after a prolonged feeding period of 9 days, we interrogated whether milk miRNAs could possibly accumulate more rapidly. Again, we utilized the preterm piglet model and tested whether and how fast bloodstream accumulation of xenomiRs as tracers after the initiation of enteral feeding and after GI transit takes place. Therefore, a dietary colostrum bolus was supplemented with reporter miRNAs from *Caenorhabditis elegans* (cel-miR-39-5p and cel-miR-39-3p [Figure 3C]). For those reporter miRNAs as well as for endogenous control miRNAs (miR-148a-3p, miR-143-3p, miR-99b-5p) and U6 snRNA, we analyzed their differential enrichment in fractionated EVs using ExoQuick exosome precipitation reagent (SBI System Biosciences) and qPCR assessment of purified small RNAs. As suggested by microvolume electrophoresis (Figure 3A top) and PAGE (Figure 3A bottom), the yield of small RNAs from the EV fraction was comparatively higher than from the milk serum supernatant from human, bovine, and porcine colostrum. In qPCR analyses, using the endogenous controls for all human, bovine, and porcine colostrum samples, miR-143-3p and U6 snRNA were detectable in both the EV fraction and the supernatant (Supplemental Figure 8A). In contrast, we observed miR-99b-5p only in the EV fraction because melting curve analysis for miR-99b-5p suggested (partial) nonspecific amplification. In all cases, we also observed that CT values for EV-associated amplicons were significantly smaller than for amplicon from the supernatant. We then tested whether the spiked cel-miRs were associated with EVs. To do this, we analyzed the small RNAs recovered from the EV fraction by qPCR and compared the samples with the serial cel-miR-395p/-3p dilutions with non-template-control (NTC) samples that did not contain cel-miR-39-5p/3p (Figure 3B, Supplemental Figure 8B,C). For all human, bovine, and porcine colostrum samples, similar results showed that the endogenous controls (miR-99b-5p, miR-143-3p, miR-148a-3p, and U6 snRNA) were largely uniformly and specifically detectable. Both cel-miR-39-5p and also cel-miR-39-3p were specifically and evidently detectable depending on the serial dilution. The experiments thus allow us to conclude that the miRNA tracer introduced into the colostrum samples co-precipitated with the EV fraction in remarkable amounts.

**Figure 3.**
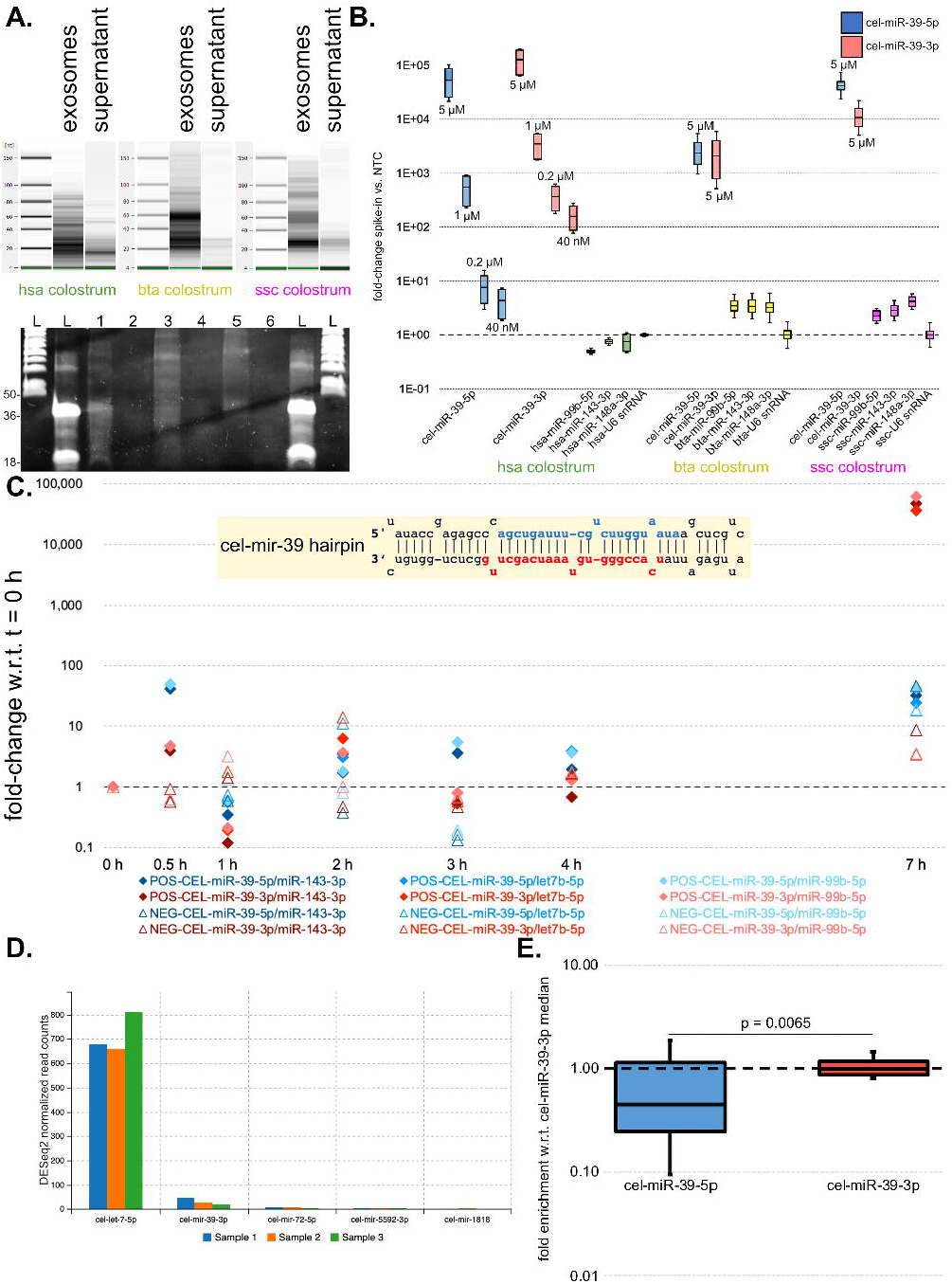
Reporter cel-miR-39-3p accumulates in the bloodstream and cel-miR-39-5p/-3p co-precipitate with Argonaute/AGO2 from homogenized intestinal biopsies. **A.** Small RNAs were analyzed via microvolume electrophoresis using a DNA 1000 chip and the Agilent Bioanalyzer 2100 (top) as well as PAGE (bottom). Abbreviations: L (ladder), 1 (hsa exosomes), 2 (has supernatant), 3 (bta exosomes), 4 (bta supernatant), 5 (ssc exosomes), 6 (ssc supernatant). **B.** Results of assessment of spiked cel-miR-39-5p/-3p in the exosome fractions of human, bovine or porcine colostrum via qPCR. The y-axis shows the relative fold-change enrichment of spiked cel-miRs or endogenous control miRs versus NTC as determined by the ΔΔCT method. U6 snRNA was used for normalization. Importantly, because the amount of cel-miR-39-5p/3p in the NTC is zero, it is not possible to calculate the enrichment by fold change with conventional methods, as dividing by zero is undefined. As an approach, we therefore considered the CT values corresponding to the unspecific cel-miR-39-5p/-3p amplification as pseudo counts. **C.** Results of qPCR to trace the cel-miR-39-5p/3p reporter in the bloodstream. The yellow shaded infobox within the chart illustrates the *C. elegans* (cel)-mir-39 hairpin. Prospective guide and passenger strand sequences, which were source for spiked in reporter miRNAs, are highlighted for cel-miR-39-5p (blue) and cel-miR-39-3p (red). Accordingly, these colors were used for the qPCR amplification data points of the detected cel-miR-39-5p/3p-amplicons (below) from blood serum samples. Normalization of cel-miR-39-5p and cel-miR-39-3p was performed using endogenous pan-miR-143-3p, pan-miR-99b-5p or pan-let-7b-5p, respectively. Blue rhomboid data points: cel-miR-39-5p (POS; cel-miR-39 feeding); red rhomboid data points: cel-miR-39-3p (POS; cel-miR-39 feeding); triangular data points: (NEG; no cel-miR-39 supplementation). **D.** Using Chimira (43) for analyses of small RNA sequencing raw data from n=3 piglet blood serum miRNA-libraries and selecting ‘*C. elegans*’ as species option, we identified cel-miR-39-3p (avg. 95 raw read counts). For comparison, matching cel-let-7-5p (avg. 2145 raw read counts, sequence identity to ssc-let-7a-5p [nt 1-22]) and cel-miR-72-5p (avg. 22 raw read counts, probably being identified as ssc-miR-31-5p [1nt mismatch]) are shown as well as ambiguous matches for cel-miR-5592-3p (avg. 17 raw read counts) and cel-miR-1818 (avg. 5 raw read counts). Notably, the charts y-axis is DESeq2 normalized read counts. **E.** Qualitative evidence for the presence of cel-miR-39-5p and cel-miR-39-3p in AGO2 immunocomplexes was achieved, when monoclonal mouse pan-argonaute/AGO2 anibodies (Sigma-Aldrich, MABE56, clone 2A8 (46)) were used for pull-down experiments from n=3 intestinal tissue samples. Prior to qPCR miRNA cDNA libraries were prepared from co-precipitated small RNAs. The chart exhibits quantitative differences between pulled-down cel-miR-39-5p and cel-miR-39-3p. Relative fold-changes with respect to the cel-miR-39-3p median were determined using the ΔΔCt method and miR-143-3p as well as miR-99b-5p for normalization.

At different time points post-feeding (t_0_=0 min, t_1_=30 min, t_2_=1 h, t_3_=2 h, t_4_=3 h, t_5_=5 h and t_6_=7 h) peripheral blood was sampled. Then total serum RNA was used for miRNA library preparation and successive analyses of enrichment of both reporter cel-miRNAs by qPCR. For confirmation, miRNA-seq was conducted from serum sampled 7 h post-feeding. Along most time points of sampling, we did not observe qPCR signals above threshold, but 7 h post feeding we obtained a positive qPCR signal for cel-miR-39-3p (Figure 3C). The positive qPCR signal for cel-miR-39-3p was verified by miRNA-seq (Figure 3D). At the same time point no qPCR signal above threshold was observable for cel-miR-39-5p.

### Dietary supplemented cel-miR-39-5p and cel-miR-39-3p co-precipitated with AGO2

The biological activity of miRNAs requires their loading into AGO2-containing protein complexes. Thus, we analyzed whether cel-miR-39-5p/-3p miRNAs can be pulled-down from piglet intestinal biopsies 7h post-feeding using anti-pan-AGO2 monoclonal antibodies (Sigma-Aldrich, MABE56, clone 2A8). Via qPCR, both cel-miR-39-5p as well as cel-miR-39-3p could be detected in AGO2-precipitates from pre-term piglets, which were fed with cel-miR-39-5p/-3p-supplemented colostrum as described above (Figure 3E).

### Weak evidence for DDX3 targeting through bta-mir-677 in HIEC-6 cells was superimposed by several probable false-positive predictions of miRNA-mRNA interactions

Using adenovirus type 5 (Ad5)-based miRNA gene transfer, we studied exemplarily the potential influence of bta-mir-677 (and other bta-miRs, see below) on bioinformatically predicted human targets in primary fetal human intestinal epithelial cells (HIEC-6) (53). By utilizing this transduction method, we efficiently overcame the typically disadvantageous signal-to-noise ratios of standard transfection protocols (Supplemental Figure 5). Transcriptome profiling of untreated and Ad5-treated cells confirmed that this model well fulfilled important criteria for the purpose of miRNA experiments, because the Ad5-treatment apparently did not lead to significant changes in the HIEC-6 transcriptome profiles: Under primary cell culture conditions and under the influence of Ad5-mediated miRNA transfer HIEC-6 retained fetal epithelial cell characteristics, and important gene activities related to miRNA biogenesis and RNAi activity were observable (Figure 4A).

**Figure 4.**
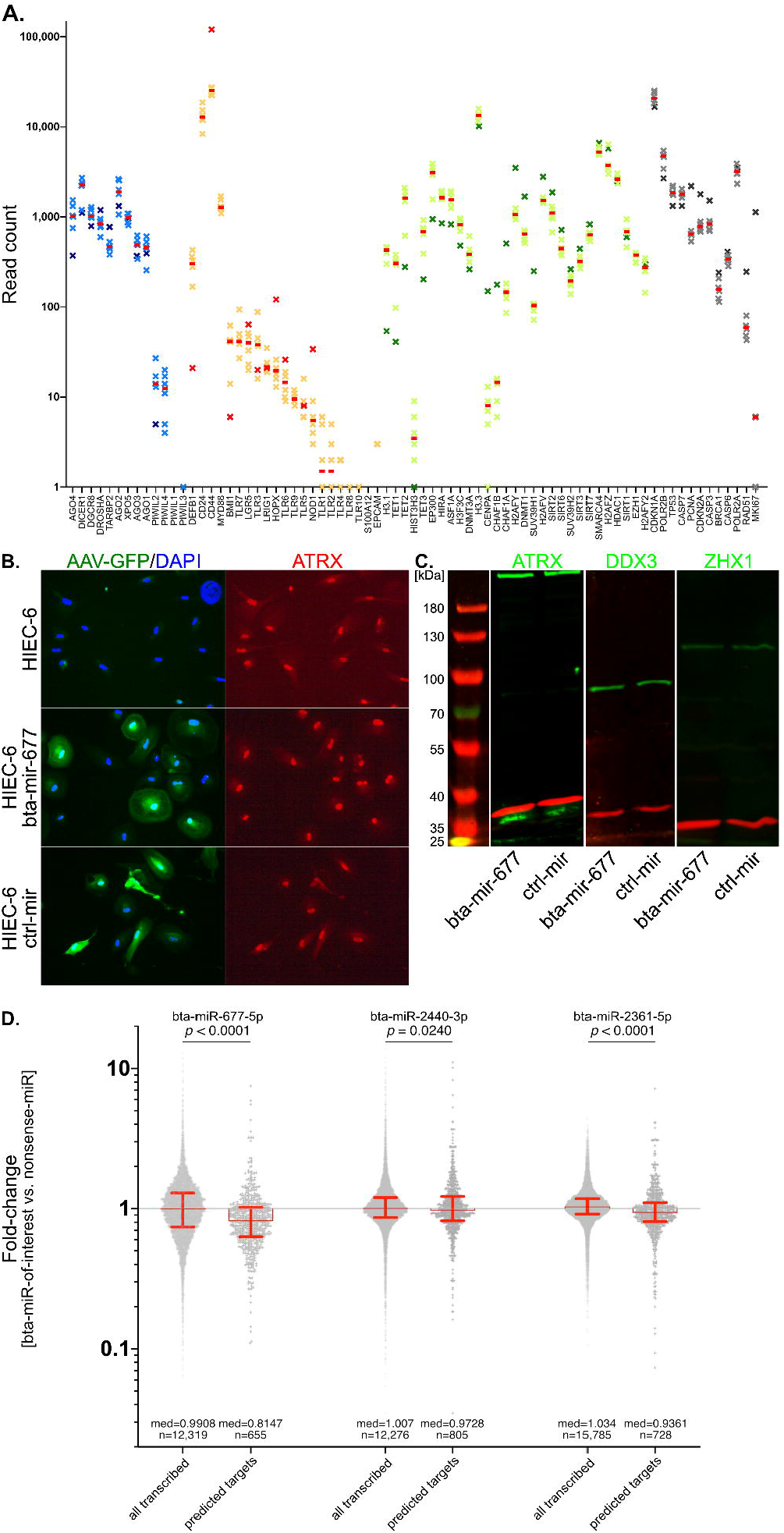
Tests for the suppression of predicted human mRNA targets of bta-miR-677-5p on the protein level for selected candidates and for 4 bta-miRs on the whole transcriptome level. **A.** Transcription patterns are shown for selected genes, which are markers for primary fetal epithelial cells or selected cellular processes. Our goal was to rule out that Ad5 treatment could induce massive off-target effects, such as dedifferentiation, changes in epigenome plasticity, changes in proliferation, loss of parts of the RNAi machinery. Mapped raw read counts reflect HIEC-6 gene expression profiles (untreated HIEC-6 and HIEC-6, which underwent Ad5-mediated miRNA gene transfer, i.e. nonsense miR, bta-miR-677-5p, bta-miR-2400-5p, bta-miR-2440-3p, bta-miR-2361-5p). Selected groups of genes are shown: gene expression related to miRNA biogenesis and RNAi activity (blue asterisks), intestinal epithelial cell markers and innate immunity gene expression (orange asterisks); DNA methylation and chromatin structure related gene expression (green asterisks); cell cycle and proliferation related gene expression (grey asterisk). Untreated HIEC-6 are highlighted in each data column through darker colored asterisks. Each red line marks the median read count for each data column. **B.** Immunofluorescence microscopy revealed no differences in the nuclear localization of ATRX upon Ad5-mediated bta-miR-677 gene transfer into primary HIEC-6 cells. ATRX predominantly exhibited nuclear localization and therein we observed focal accumulations in wt-HIEC-6 as well as in bta-mir-677-treated HIEC-6 and ctrl-mir-treated HIEC-6. We routinely focused our view to areas wherein cells exhibited GFP-signals of different intensity. This should enable us to detect differences in ATRX distribution, which hypothetically could depend on the level of expressed bta-mir-677. Again, no differences were seen between treated cells and untreated controls. Notably, for PLCXD3 no reliable signals were observed in HIEC-6 cells at all. **C.** When normalized with the GAPDH, no differences in the abundances of ATRX or ZHX1 were observed upon Ad5-mediated bta-miR-677 gene transfer into primary HIEC-6 cells in Western analyses, when compared to wild-type (wt) HIEC-6 (not shown) or HIEC-6 expressing a control-mir. Using two replicative measurements for quantitative Western Blot analyses via LI-COR Empiria Studio Software, we measured a 24.45% (SD: ± 1.68%) DDX3 signal reduction in bta-mir-677-treated vs. wt and, respectively, a 37.90% (SD: ± 1.90%) DDX3 signal reduction in bta-mir-treated vs. ctrl-mir-treated HIEC-6. **D.** Effects of Ad5-mediated expression of 3 selected bta-miRs were studied in HIEC-6 (bta-miR-677-5p; bta-miR-2440-3p; bta-miR-2361-5p) in comparison to HIEC-6 expressing a Ad5-mediated nonsense-miR. Relative fold-changes were compared for the whole HIEC-6 transcriptome and the lists of expressed predicted targets. Predicted targets for each bta-miR comprised of hundreds of mRNAs (i.e. bta-miR-677-5p [n=655]; bta-miR-2440-3p [n=805]; bta-miR-2361-5p [n=728]). Statistical power calculations were done using the nonparametric Wilcoxon–Mann–Whitney method and GraphPad Prism 8.4.3 software.

For bta-miR-677-5p, miRDB (38) output a list of 655 predicted human target mRNAs. From this list, we selected ATRX, DDX3, PLCXD3 and ZHX1 for analyses by immunofluorescence microscopy and quantitative Western blots (Figure 4B,C). These predicted targets had high miRDB scores, and they were annotated being potential master regulators of other gene activities. However, from this unavoidably arbitrarily selection, none of the candidates exhibited differences in the quantity or subcellular distribution using microscopy (Figure 4B). On the scale of whole cell populations, we conducted quantitative Western blot analyses. Interestingly, for DDX3, analyses of two Western blots revealed a 24.45% reduction (SD: ± 1.68%) of DDX3 signals in bta-mir-677-treated vs. wild-type and a 37.90% (SD: ± 1.90%) DDX3 signal reduction in bta-mir-treated vs. ctrl-mir-treated HIEC-6. Notably, the effects were not visible by eye (Figure 4C).

Since the above methods of experimental target validation on the protein level relies on a relatively arbitrary selection of putative targets and laborious non-holistic validation, we additionally analyzed the influence of Ad5-mediated miRNA gene transfer using a transcriptomics (mRNA-seq) approach to study the potential influence of three different bovine miRNAs-of-interest (miRoI) (bta-miR-677-5p, bta-miR-2440-3p and bta-miR-2361). Through normalization (miRoI-treated vs. nonsense-miR-treated HIEC-6) we obtained fold-changes in gene expression for every transcribed mRNAs. We then compared the fold-changes of all expressed predicted target mRNA with the entire transcriptome of nonsense-miR-treated HIEC-6 (Figure 4D). Whereas most genes’ transcripts remained relatively unaffected by miRoI-treatment (fold change: approx. 1.0), we observed an enrichment of significantly downregulated genes (fold change: <1.0) within the group of the predicted miRoI targets (Median fold-changes all transcripts/predicted targets: bta-miR-677-5p: 0.9908/0.8147 [*p* < 0.0001]; bta-miR-2440-3p: 1.0070/0.9728 [*p* = 0.0240]; bta-miR-2361-5p: 1.0340/0.9361 [*p* < 0.0001]). Notably, we did not see any correlation between the degree of observed downregulation and the individual position/score as a result of target predictions.

## Discussion

One exceptional previous study in the field used an elegant approach looking at the intestinal uptake of milk miRNAs in mmu-mir-375-knockout and mmu-mir-200c/141-knockout mice offspring, which received milk from wild-type foster mothers. Here, no evidence for intestinal epithelial uptake of miRNA and/or its enrichment in blood, liver and spleen was seen (27). Contradicting results were obtained from a more recent study that simulated infant gut digestion *in vitro* and analyzed the uptake of human milk miRNAs by human intestinal epithelial crypt-like cells (HIEC) deriving from fetal intestines at mid-gestation. Here, milk miRNA profiles remained stable after digestion and evidence for the uptake of milk-derived exosomes in HIEC was reported (28).

We can provide more clarity in this regard, since the discovery of many taxon-specific miRNAs allowed the conclusion that dietary xenomiRs from foreign species’ milks could be exploited as molecular tracers *in vivo* to verify their gastrointestinal (GI) passage as well as systemic and cellular uptake in human or animal models. In human newborns dietary-derived bta-miRs survived the gastrointestinal transit. This demonstrated that nutritional uptake of milk miRNAs leads to their intestinal enrichment and suggests their stable sequence integrity during gastrointestinal (GI) transit in human preterm neonates.

In preterm piglets our initial observation was that cattle-specific miRNAs became enriched in intestinal cells after prolonged (9-days) enteral feeding with bovine colostrum or formula supplementation. Thereby, bta-miR-2904 occupied a high rank among the most abundant miRNAs within intestinal cells. This suggested that a dietary miRNA could in principle reach biologically significant levels if being taken up by cells. Remarkably, for many identical miRNAs present in both bovine milk and porcine intestinal cells, we could not distinguish where they originated, i.e., whether they came from food or were produced by intestinal cells or both. It is also notable that none of the interrogated bta-miRs was detected in cerebrospinal fluid samples from piglets receiving enteral feeding of bovine colostrum (52), suggesting their limited blood-to-cerebrospinal fluid permeability or, respectively, indicating that they did not reach detectable concentrations.

If breast milk miRNAs were to fulfill signaling functions within a hypothetical, discrete perinatal ‘window of opportunity’ that opens during the introduction of enteral feeding, a much more rapid accumulation of miRNAs would be required. We assume that meanwhile this window is open, the intestine develops as an environment-organism interface to coordinate microbiome homeostasis. Therefore, we considered that intestinal immaturity in preterm neonates could provide a clinically significant niche for (xenobiotic) milk miRNA influence. To investigate this, we performed additional xenomiR-tracer feeding experiments in neonatal preterm piglets for a period of 7 h after the onset of enteral feeding. We spiked human, bovine or porcine colostrum with cel-miR-39-5p and cel-miR-39-3p, and this led to their effective equilibration with the EV fraction. We assumed that externally supplied miRNAs were linked to endogenous miRNAs found in extracellular vesicles (EVs), which are believed to be bioavailable and non-nutritional. However, we caution that our experiments are insufficient to draw definitive conclusions whether and how spiked miRNAs could become packaged inside EVs, including whether specific EV subtypes such as exosomes are involved (see above: Statement on methodological limitations). Feeding with cel-miR-39-5p/-3p-supplemented colostrum bolus led to bloodstream enrichment of cel-miR-39-3p. From our qPCR results, we concluded that a transit of considerable amounts of cel-miR-39-3p from the GI tract into the bloodstream took place, whereas cel-miR-39-5p was not detected within the observation period. The timing of reporter miRNA accumulation in serum roughly appeared to be in agreement with GI transit times of radiotracers in suckling piglets (21 days old; approx. 1.5-3.5 h until 75% of gastric emptying and >9.5 h until 75% intestinal emptying) [28].

Importantly, we observed AGO2-loading with both cel-miR-39-5p and cel-miR-39-3p in intestinal cells. This suggested that AGO2 in intestinal cells was loaded with both the guide and the passenger strands, which resulted from processed cel-mir-39 (Figure 3E) and that AGO2-loaded milk miRNAs could indeed act as regulators in the development of gut homeostasis or other developmental processes, which begin perinatally or early postnatally. Due to the presence of cel-miR-39-5p in AGO2-immunocomplexes, we assume that its bloodstream enrichment could simply lag behind cel-miR-39-3p instead of being selectively excluded. However, the mechanistic details of miRNA transport and selection criteria for AGO2-loading remain unknown so far.

Taken together, our experimental trajectory suggests that dietary uptake of milk miRNAs can lead to their loading onto AGO2 in neonatal intestinal epithelial cells. Thus, vertical transmission of maternal milk miRNA signals via GI transit to responsive cells and systemic bloodstream distribution is apparently possible. This is in agreement with the ‘functional hypothesis’ but contradicts the ‘non-functional’ hypothesis that milk miRNAs could solely serve as a nucleoside source for the newborn (54). Our tracer experiments provide good experimental justification to address superordinate questions about milk miRNA-induced PTGS in neonates. However, key questions remain unanswered: we have no clear idea which of the hundreds of different miRNAs found in milk, with different abundances, could be important and how selective loading of AGO2 in milk miRNA-responsive cells could work. It is possible that only some specific miRNAs find specific mRNA duplex partners, depending on their presence in specific cell types and transcriptional timing. An important future concept to look into further will be the exact mechanisms of milk miRNA packaging, transport and uptake by recipient cells via EVs. Systemically, it remains unknown how far-reaching and selective milk miRNAs’ influence could be. Could biologically relevant miRNA accumulation be restricted to the intestine, or does their observation in the bloodstream indicate a systemic distribution to other target tissues/cells? As important as the place of milk miRNA activity is the time of action during ontogeny. The possibly most convincing period could be during a relatively narrow ‘window of opportunity’ that could open perinatally and persist during the introduction of enteral feeding, when the infant’s intestine develops as an environment-organism interface to coordinate microbiome homeostasis.

If anticipating a vertical transmission of PTGS via milk miRNAs the superordinate problem is, which mRNAs are true targets. In this work, we have found weak evidence for DDX3 targeting through bta-mir-677 in HIEC-6 cells. The underlying experiments relied on a mRNA target prediction algorithm, whose results were probably superimposed by several false-positive predictions of miRNA-mRNA interactions. Clearly, target prediction depending mostly of 5’-seed complementarity remains leaky for simple reasons: First, because searches for sequence matches in mRNA databases usually cannot consider whether a potential target is transcribed in a cell or tissue-of-interest or not. Second, prediction algorithms do not consider that within a cell different miRNAs and mRNAs compete for being loaded to a limited number of Argonaute (Ago) proteins [29]. Moreover, given that mRNA decay and blocking of translation exist as alternative miRNA interference mechanisms, it is not clear whether PTGS can be detectable directly on the mRNA level and/or the protein level only. Consequently, the identification of valid miRNA-target interactions and their influence on gene regulatory networks requires detailed experimental analyses. These results suggested that bta-mir-677 expression in HIEC-6 cells could indeed influence the expression of one of the predicted human mRNA targets, leading to reduced translated DDX3 protein. Even though the latter observation could give some hint for an influence of bta-miR-677-5p in HIEC-6 cells, the relatively arbitrary putative target selection and laborious validation is unrewarding and non-holistic. In summary, we have efficiently applied Ad5-mediated gene transfer in HIEC-6, establishing continuous, but probably non-natural cellular levels of ectopically expressed miRNAs. This might lead to enforced AGO2-loading caused by outnumbered availability of the substrate (dog-eat-dog effect). These conditions may not fully reflect the in vivo situation in the neonate intestine, but nevertheless might be useful to study the influence of (xenobiotic) milk miRNAs on gene regulatory networks. This approach could plausibly help to explain the observed increase of severe intestinal complications such as NEC in premature infants fed with bovine based formula milk. However, we clearly emphasize that postulating potential harmful influence would be an overstated conclusion from our experiments. But for the first time in human history, the newest developments in intensive care neonatology allow the survival of extremely vulnerable preterm neonates, whereby severe complication are often associated with incomplete intestinal development. Thus, if the ‘window of opportunity’ hypothesis discussed above holds, the consideration of milk xenomiRs’ side effects on early preterm and term neonatal development seems well justified and would encourage advanced future studies.

Finally, we conclude:

- Dietary-derived miRNAs survived gastrointestinal transit in human newborns, demonstrating the intestinal enrichment of milk miRNAs.
- In preterm piglets, cattle-specific miRNAs became enriched in intestinal cells after enteral feeding with bovine colostrum or formula.
- Feeding with cel-miR-39-5p/-3p-supplemented colostrum in neonatal preterm piglets led to bloodstream enrichment of cel-miR-39-3p.
- Dietary uptake of milk miRNAs can lead to AGO2 loading in neonatal intestinal epithelial cells, suggesting vertical transmission of maternal milk miRNA signals.
- We provided experimental justification for further research into milk miRNA-induced post-transcriptional gene silencing in neonates.
- Questions remain regarding which miRNAs are important and how selective AGO2 loading in milk miRNA-responsive cells works, as well as the systemic influence of milk miRNAs.

## Declarations and ethics statement

### Ethics approval

This study was conducted with approval of the Witten/Herdecke University Ethics board (No. 41/2018). A participant flowchart for human and animal milk specimen collection is provided as Supplemental Figure 2. All animal procedures were approved by the National Council on Animal Experimentation in Denmark (protocol number 2012-15-293400193). A participant flowchart for piglet intestinal specimen collection is provided as Supplemental Figure 3. The authors state that they have obtained appropriate institutional review board approval or have followed the principles outlined in the Declaration of Helsinki for all human or animal experimental investigations. In addition, for investigations involving human subjects, informed consent has been obtained from all participants. Patients or the public were not involved in the design, or conduct, or reporting, or dissemination plans of our research.

### Informed consent statement

Informed written consent was obtained by all involved donors.

### Data availability statement

The datasets generated and/or analyzed during the current study are available in the NCBI BioProject repository as fastq.gz files of miRNA-seq and mRNA-seq (SubmissionIDs: SUB7584111/ SUB9897357/BioProject ID: PRJNA740153/URL: https://www.ncbi.nlm.nih.gov/bioproject/740153).

### Competing interests statement

All authors declare that there is no conflict of interests.

## Funding

This study was funded by the HELIOS Research Centre (JP) and the NEOMUNE research program (PTS) from the Danish Research Council. University of Copenhagen has filed a patent concerning the use of bovine colostrum for groups of pediatric patients. The authors have no other relevant affiliations or financial involvement with any organization or entity with a financial interest in or financial conflict with the subject matter or materials discussed in the manuscript apart from those disclosed. No writing assistance was utilized in the production of this manuscript.

## Data Availability

The datasets generated and/or analysed during the current study are available in the NCBI BioProject repository as fastq.gz files of microRNA-seq and mRNA-seq (SubmissionIDs: SUB7584111/ SUB9897357/BioProject ID: PRJNA740153/URL: https://www.ncbi.nlm.nih.gov/bioproject/740153).

## Acknowledgements

We gratefully acknowledge Gilles Gasparoni (Institut für Genetik/Epigenetik, Universität des Saarlandes, Germany) to enable and support our mRNA deep sequencing analyses under hard conditions during the first European peak period of the SARS-CoV-2 pandemic. We further thank Dr. Arne Lawrenz (director of Wuppertal Zoo) for providing Okapi (Lomela’s) milk as well as Stefan König (Hof Sackern, Wetter (Ruhr), Germany) for donating bovine colostrum.

Authors’ responsibilities were as follows: JP and PPW designed the study and wrote the paper. SR conducted Ad5-mediated miRNA gene transfer experiments under supervision and with the help of FK, FJ and CH. Further, SR prepared mRNA-seq libraries. FK and JS developed the Ad5-mediated miRNA gene transfer. CAH traced miRNAs in piglet blood and intestine and human stool. MA, AP and VO performed luciferase experiments. Animal treatments as well as sample collection and preparation were done by DNN, PPJ and PTS. PPW, FG, TT, MA, ACWJ and SW contributed to sampling of human and animal milk specimens. PPW, FG, VW and CAH generated RNA libraries, conducted qPCR experiments and NGS. JP and CAH performed Argonaute pull-down experiments. JP and PPW supervised undergraduates and graduate students. JP supervised the study, analyzed the data and wrote the manuscript with the help of PPW, ACWJ, CH and FK. All authors have read and approved the final manuscript.

## Notes

### Competing Interest Statement

The authors have declared no competing interest.

### Clinical Trial

Not relevant

### Funding Statement

This study was funded by the HELIOS Research Centre (JP) and the NEOMUNE research program (PTS) of the Danish Research Councils. University of Copenhagen has filed a patent concerning the use of bovine colostrum for groups of paediatric patients. The authors have no other relevant affiliations or financial involvement with any organisation or entity with a financial interest in or financial conflict with the subject matter or materials discussed in the manuscript apart from those disclosed. No writing assistance was utilised in the production of this manuscript.

### Author Declarations

Ethics approval and consent to participate This study was conducted with approval of the Witten/Herdecke University Ethics board (No. 41/2018). All animal procedures were approved by the National Council on Animal Experimentation in Denmark (protocol number 2012-15-293400193). The authors state that they have obtained appropriate institutional review board approval or have followed the principles outlined in the Declaration of Helsinki for all human or animal experimental investigations. In addition, for investigations involving human subjects, informed consent has been obtained from all participants.

### Summary of Updates

-Added experiment: Precipitation and analyses of spike miRs in extracellular particles from colostrum specimens. Added description in Materials and Methods section. Results considered in Figure 3. Results and Discussion. Some previous content of Figure 3 is now in the Supplementary Data. -Language editing in Abstract, Introduction, Results and Discussion. -Graphical Abstract was removed. -Reformatted article according to AJCN style. -Updated Supplementary Files.

