## Supplemental Table 1 for "Uncovering the gastrointestinal passage, intestinal epithelial cellular uptake and AGO2 loading of milk miRNAs in neonates using xenomiRs as tracers"

**Uncovering the gastrointestinal passage, intestinal epithelial cellular uptake and AGO2 loading of milk miRNAs in neonates using xenomiRs as tracers**

Patrick Philipp Weil^1^, Susanna Reincke^1^, Christian Alexander Hirsch^1^, Federica Giachero^1^, Malik Aydin^1,2^, Jonas Scholz^3^, Franziska Jönsson^3^, Claudia Hagedorn^3^, Duc Ninh Nguyen^4^, Thomas Thymann^4^, Anton Pembaur^1^, Valerie Orth^5^, Victoria Wünsche^1^, Ping-Ping Jiang^4,6^ Stefan Wirth^2^, Andreas C. W. Jenke^7^, Per Torp Sangild^4^, Florian Kreppel^3^, Jan Postberg^1,*^

*corresponding author

^1^Clinical Molecular Genetics and Epigenetics, Faculty of Health, Centre for Biomedical Education & Research (ZBAF), Witten/Herdecke University, Alfred-Herrhausen-Str. 50, 58448 Witten, Germany

^2^HELIOS University Hospital Wuppertal, Children’s Hospital, Centre for Clinical & Translational Research (CCTR), Witten/Herdecke University, Heusnerstr. 40, 42283 Wuppertal, Germany

^3^Chair of Biochemistry and Molecular Medicine, Faculty of Health, Centre for Biomedical Education and Research (ZBAF), Witten/Herdecke University, Stockumer Str. 10, 58453 Witten, Germany

^4^Comparative Pediatrics and Nutrition, Department of Veterinary and Animal Sciences, University of Copenhagen, Copenhagen, Denmark

^5^HELIOS University Hospital Wuppertal, Department of Surgery II, Centre for Clinical & Translational Research (CCTR), Witten/Herdecke University, Heusnerstr. 40, 42283 Wuppertal, Germany

^6^School of Public Health, Sun Yat-sen University, Guangzhou, China

^7^Klinikum Kassel, Zentrum für Kinder- und Jugendmedizin, Neonatologie und allgemeine Pädiatrie, Mönchebergstr. 41-43, 34125 Kassel, Germany

| **formula/powdered milk product** | **manufacturer** |
| --- | --- |
| Enfalac A+ | Mead Johnson Nutrition – Chicago, USA |
| Enfamil Human Milk Fortifier | Mead Johnson Nutrition – Chicago, USA |
| Aptamil Pronutra ADVANCE PRE | Milupa Nutricia GmbH – Frankfurt am Main, GER |
| Aptamil PDF | Milupa Nutricia GmbH – Frankfurt am Main, GER |
| NutriniDrink MultiFibre | Nutricia GmbH – Erlangen GER |
| Nutrilon Pronutra | Nutricia GmbH – Erlangen GER |
| Protifar Neutral | Nutricia GmbH – Erlangen GER |
| SERAVIT | Nutricia GmbH – Erlangen GER |
| BEBA PRO HA PRE | Nestlé S.A. – Vevey, CH |
| PRE NAN FM 85 | Nestlé S.A. – Vevey, CH |
| Skim Milk Powder | Fluka Analytical - Seelze (D) |

**A.** List of formula/powdered milk products used in this study.

| **oligo name** | **Sequence (5’→ 3’)** | **note** |
| --- | --- | --- |
| bta-miR-2478-3p-fw | ttccatggtgtcagaagtggga | DNA |
| bta-miR-2478-5p-fw | ttccagctcaccaccgaccccc | DNA |
| bta-miR-2887-3p-fw | ttccacgcgccccgtgtcccgg | DNA |
| bta-miR-2887-5p-fw | ttccacgcaccggaccccggtc | DNA |
| bta-miR-2904-3p-fw | ttccaggggacggcggggga | DNA |
| bta-miR-2904-5p-fw | ttccagaggccaaccgaggctc | DNA |
| bta-miR-677-3p-fw | ttccacaaacgacttcggtcta | DNA |
| bta-miR-677-5p-fw | ttccagtcagaagctgctcatc | DNA |
| bta-mir-677-hairpin | cucacugaugagcagcuucugacacagugaagcugcucaucagugaguuuuu | RNA |
| bta-miR-677_target_sense | catgcagacaatcagtgaagtccaattag | DNA |
| bta-miR-677_target_asense | ctagctaattggacttcactgattgtctgcatgagct | DNA |
| cel-miR-39-3p | ucaccggguguaaaucagcuug | RNA |
| cel-miR-39-3p_lib_fw | ccgagaattccacaagctgatt | DNA |
| cel-miR-39-3p_lib_rv | tacagtccgacgatctcacc | DNA |
| cel-miR-39-3p_fw | tgatttacacccggtgactagca | DNA |
| cel-miR-39-5p | agcugauuucgucuugguaaua | RNA |
| cel-miR-39-5p_lib_fw | ccgagaattccatattaccaag | DNA |
| cel-miR-39-5p_lib_rv | tacagtccgacgatcagctg | DNA |
| cel-miR-39-5p_fw | ccaagacgaaatcagctctagca | DNA |
| library_P1_miR_rev | aatgatacggcgaccaccgag | DNA |
| library_P2_miR_rev | caagcagaagacggcatacgag | DNA |
| cel-miR-39-3p-fw | tcaccgggtgtaaatcagcttg | DNA |
| cel-miR-39-5p-fw | agctgatttcgtcttggtaata | DNA |
| miR-148a-3p-fw | tcagtgcactacagaactttgt | DNA |
| miR-143-3p-fw | tgagatgaagcactgtagctc | DNA |
| miR-99b-5p-fw | cacccgtagaaccgaccttgcg | DNA |
| U6 snRNA | Mir-X™ miRNA First Strand Synthesis Kit (Takara) | DNA |
| miR-143-3p_lib_fw | ccgagaattccagagctacagt | DNA |
| miR-143-3p_lib_rv | tacagtccgacgatctgaga | DNA |
| (pan)-miR-143-3p_fw | ttccagagctacagtgcttca | DNA |
| miR-99b-5p_lib_fw | ccgagaattccacgcaaggtcg | DNA |
| miR-99b-5p_lib_rv | tacagtccgacgatccaccc | DNA |
| (pan)-miR-99b-5p_fw | ttccacgcaaggtcggttctac | DNA |

**B.** Oligonucleotides used in this study
